# Depression Risk and Work Hours in Training Physicians Before and During the COVID-19 Pandemic

**DOI:** 10.1101/2025.03.09.25323517

**Authors:** Jennifer L. Cleary, Karina Pereira-Lima, Xianda Ma, Lihong Chen, Margit Burmeister, Lisa M. Meeks, Zhuo Zhao, Jun Ye, Yu Fang, Zhenke Wu, Elena Frank, Ruyuan Zhang, Suhua Zeng, Qian Zhao, Douglas A. Mata, Amy Bohnert, Weidong Li, Srijan Sen

## Abstract

**Importance:** In the general population, depression increased with the onset of the COVID-19 pandemic. In addition to the general pandemic impact, training physicians faced many sudden and dramatic changes in their training environment. However, the effects of these changes on the mental health of training physicians remains unknown.

**Objective:** To identify change in depression risk among training physicians with the onset of the COVID-19 pandemic and factors associated with risk.

**Design:** Prospective cohort study.

**Setting:** University- and community-based health care institutions in the United States and Shanghai, China

**Participants:** First-year resident physicians (interns) serving during the 2018-19 (n=1844), 2019-20 (n=1201), and 2020-21 (n=2448) academic years (U.S. sample); interns serving during the 2021-22 academic year (n=471) (Shanghai sample)

**Main Outcomes and Measurements:** Depressive symptoms (Patient Health Questionnaire-9 [PHQ-9]) and work hours were assessed quarterly for all U.S. cohorts. The 2019-20 cohort completed supplemental surveys of these measures in April and May 2020. Shanghai sample interns were assessed for depressive symptoms (PHQ-9) and work hours quarterly before, during, and after the 2022 lockdown.

**Results:** Within the 2019-20 U.S. cohort, depressive symptom scores decreased from the pre-pandemic (September, December) to the pandemic period (April, May, June) (5.5 [3.9] vs. 4.9 [4.3], *p*<0.001). In causal mediation analysis, 62% of this change was mediated through work hours (0.62, 95% CI [0.44-1.00]). Descriptive comparisons of this cohort with cohorts training immediately before (2018-19) and after (2020-21) the pandemic onset demonstrated that both work hours and depressive symptoms were significantly lower in spring 2020, but returned to pre-pandemic levels by fall 2020. In the parallel Shanghai cohort serving during the April 2022 lockdown, we found a similar magnitude drop in depressive symptoms (5.6 [3.3] vs. 4.9 [4.8], p=0.005), with 64% of the effect mediated through work hours (0.64, 95% CI [0.24-1.84]).

**Conclusions and Relevance:** Interns experienced a 11% decrease in depressive symptoms with the onset of the pandemic, which was primarily driven by reduced work hours. The identified associations between work hours and depressive symptoms early in the pandemic may inform strategies to support physician wellness moving forward.

**KEY POINTS:** *Question:* How did depressive symptoms in training physicians change during the COVID-19 pandemic?

*Findings:* In a prospective cohort study including 1201 U.S. first-year residents during the 2019-2020 academic year, we found an 11% decrease in depressive symptoms with the pandemic onset. Causal mediation analysis identified that work hours mediated 62% of this change. These findings were replicated in a parallel cohort of 471 Shanghai residents, where work hours mediated 64% of the 11% decrease in depressive symptoms during the 2022 lockdown.

*Meaning:* The identified decreases in depressive symptoms mediated through work hours can inform future reforms to support resident well-being.

## INTRODUCTION

Physician residency training is a time of high stress, with 25-30% of residents screening positive for depression at any given time^1^. Across the world, the early phase of the coronavirus disease 2019 (COVID-19) pandemic posed an unprecedented public health challenge and was associated with an increase in depressive symptoms in the general population^2,3^. As observed during previous pandemics such as SARS^4^ and Ebola,^5^ working under pandemic conditions can result in adverse mental health consequences for healthcare providers^6–9^. Many institutions and residency programs rapidly adopted measures to support residents, such as increasing access to psychological support and reducing redundant work^10,11^, while regulatory agencies and payers implemented changes to reduce physician administrative burden^12^. Combined with these measures, the cancellation and postponement of non-urgent appointments and elective procedures further contributed to an overall decrease in workload for training physicians. Overall, it is unclear how the pandemic and resulting system changes collectively affected the well-being of training physicians.

In this study, we utilized two prospective cohort studies of first-year residents to assess the mental health of residents, and associated factors, before and during the COVID-19 pandemic. First, utilizing the large national Intern Health annual cohort study, we tracked depressive symptoms of first-year resident physicians (interns) in the U.S., working during the onset of the pandemic from September 2019 through June 2020, and examined the role of work hours in mediating observed changes in depressive symptoms within this cohort. Additionally, we compared the 2019-20 cohort with US cohorts who trained immediately prior to (September 2018-June 2019) and after (September 2020-June 2021) the onset of the pandemic. To assess the reproducibility of our findings, we examined the trajectory of depressive symptoms in a parallel cohort of Shanghai (China) intern physicians working before (October 2021-January 2022), during (April 2022), and after (July 2022) the primary COVID-19 lockdown in Shanghai.

## METHODS

### Study Design, Setting, and Participants

#### U.S. Sample

The Intern Health Study is a national prospective annual cohort study of mental health during the first year of residency training in the U.S. Following the national residency match in March of each year, individuals who were either graduating from medical school or entering residency at participating institutions were invited via email to take part in the study. Eligibility criteria included being an incoming first-year resident physician (i.e., intern) in U.S. residency programs offering graduate year 1 positions available immediately after completion of medical school. Data collection occurred before and during the onset of the COVID-19 pandemic, including the 2018-19, 2019-20, and 2020-21 academic years. In total, 11843 participants were invited to take part in the study and 6284 (53.1%) agreed to participate (n=2127 [48.9%] in the 2018-19 academic year, n=1685 [61.8%] in the 2019-20 academic year, and n=2472 [51.8%] in the 2020-21 academic year).

Participants were asked to complete surveys assessing demographic characteristics and depressive symptoms at baseline, as well as depressive symptoms and work hours quarterly throughout their internship year (September, December, March, June). Additionally, participants in the 2019-2020 cohort were sent supplemental surveys monthly in April and May 2020, to monitor these same variables during the early stage of the pandemic. Participating interns received $125 in compensation. The institutional review board at the University of Michigan approved the study.

#### Shanghai China Sample

To assess the reproducibility of findings from the U.S. cohort, we conducted a parallel analysis among a cohort of first-year resident physicians serving in Shanghai from 2021-22, before and during the Shanghai COVID-19 surge and lockdown. Utilizing similar protocols to the US sample, 1071 incoming interns at 12 Shanghai hospitals were invited via email to participate (11 invitations undeliverable). Of these, 631 individuals agreed to participate (59.5%) and 473 participants completed a baseline survey and at least one follow-up survey and were included in this analysis. Participants were asked to complete surveys assessing depressive symptoms and work hours at baseline and quarterly throughout the training year (October 2021, January 2022, April 2022, July 2022). Participants provided informed consent and were compensated ¥25.

Ethics committees of Shanghai Jiao Tong University and the University of Michigan approved this study.

### Measures

For all cohorts, participants completed an online baseline survey prior to starting internship (two months prior for U.S. cohorts, two weeks prior for the Shanghai cohort), including questions about their sex assigned at birth, age, race and ethnicity, and an assessment of their depressive symptoms using the 9-item, self-reported Patient Health Questionnaire (PHQ-9)^13^. The PHQ-9 is a well-validated^14^ measure that asks individuals to rate the frequency with which they experienced each of nine symptoms over the previous two weeks, which are summed to create a total score. In addition to the PHQ-9, within-internship surveys also included a question inquiring about participants’ work hours (“How many hours have you worked in the past week?”).

### Statistical analysis

To assess the change in depressive symptoms with the pandemic, we compared pre-pandemic (average of September and December) symptoms with their pandemic-era (average of April, May, and June) symptoms and hours for the 2019-2020 U.S. pandemic cohort, using Wilcoxon signed-rank tests for non-normally distributed data. Of note, the March survey for the 2019-2020 cohort was launched 8 days prior to the WHO’s declaration of a pandemic on March 11, 2020 and closed two weeks after the declaration. As this assessment spanned across the pandemic onset it was not included in analyses specific to pre-vs post-pandemic onset.

Given the established association^15^ between work hours and depressive symptoms, we assessed changes in work hours during the pandemic. Similar to the depressive symptom analyses, we compared pre-pandemic measures of work hours to pandemic measures of work hours using paired t-tests, as these data were normally or near-normally distributed. Next, to model the association between work hours and depressive symptom scores from September 2019 to June 2020, we used a linear mixed model (R package “lmer”). To estimate the total, direct, and causally-mediated effects of duty hours on depressive symptoms during the early pandemic, we used causal mediation models^16^ (R package “mediation”) including all interns who completed at least one PHQ-9 survey during this period.

Sample weighting can mitigate biases for non-representative sampling (poststratification weights) and loss to followup (attrition weights). Specifically, poststratification weighting matches the distribution of the current sample’s demographics to a target sample (here, the Association of American Medical College’s demographic data of all US interns in a given year). Attrition weighting identifies baseline characteristics associated with completion of follow up assessments, and creates a propensity score for follow up completion. Here, the poststratification and attrition weights were combined to form a total weight used in the causal mediation analyses (for additional details on weights in this sample, see Fang et al.^17^).

To better contextualize the change in work hours and depressive symptoms that occurred with the onset of the pandemic, we descriptively compared parallel assessments of depressive symptoms and work hours in three U.S. cohorts: the 2018-2019 cohort, who served before the onset of the pandemic, the 2019-2020 cohort, who experienced the onset of the pandemic, and the 2020-2021 cohort, who began internship after the pandemic had begun.

Finally, to assess the reproducibility of findings from the U.S. pandemic cohort in an international sample undergoing a similar structural shift in training environment, we replicated the analytical procedures used to assess changes in depressive symptoms and work hours within the 2019-20 U.S. cohort. Specifically, we assessed depressive symptoms via Wilcoxon signed-rank tests and work hours via paired t-tests to compare before, during, and after the April 2022 lockdown in the Shanghai sample. We then modeled the association between depressive symptoms and work hours using a linear mixed model (R package “lmer”) and estimated causal mediation effects (R package “mediation”).

All analyses were conducted in R, version 4.2.2 (R Program for Statistical Computing) with an alpha of <0 .05 for statistical significance.

## RESULTS

The 2019-2020 U.S. cohort served as the primary cohort for analysis (n = 1201; 681 [56.7%] female sex, mean [SD] age 27.6 [3.4] years). We utilized the pre-pandemic 2018-2019 U.S. cohort (n = 1844; 1056 [57.2%] female sex, mean age 27.5 [2.5] years) and the later-pandemic 2020-2021 U.S. cohorts (n = 2448; 1354 [55.3%] female sex, mean age 27.6 [2.7] years) as comparison cohorts. The 2021-22 Shanghai cohort consisted of 471 interns (309 (65.6%) female sex, mean age 25.4 [2.6] years. Participants and non-participants did not differ significantly in age, gender, or pre-internship PHQ-9 score. Characteristics of participating interns are presented in Table 1.

**Table 1.** Sample characteristics.

| Characteristic | Primary sample | Comparison samples |  | Replication sample |
| --- | --- | --- | --- | --- |
|  | 2019-2020 U.S.<br>cohort | 2018-2019 U.S.<br>cohort | 2020-2021 U.S.<br>cohort | 2021-2022 Shanghai<br>(China) cohort |
| Sex |  |  |  |  |
| Female | 681 (56.7) | 1056 (57.2) | 1354 (55.3) | 309 (65.6) |
| Male | 520 (43.3) | 788 (42.8) | 1094 (44.7) | 162 (34.2) |
| Age, mean (SD), years | 27.6 (3.4) | 27.5 (2.5) | 27.6 (2.7) | 25.4 (2.6) |
| Race and/or Ethnicity |  |  |  |  |
| Arab/Middle Eastern | 15 (1.2) | 38 (2.1) | 48 (2.0) | 0 (0.0) |
| Asian | 271 (22.6) | 420 (22.8) | 603 (24.6) | ** |
| Black or African American | 71 (5.9) | 83 (4.5) | 132 (5.4) | 0 (0.0) |
| Hispanic or Latino | 44 (3.7) | 71 (3.9) | 124 (5.1) | 0 (0.0) |
| Multiracial | 97 (8.1) | 161 (8.7) | 230 (9.4) | 0 (0.0) |
| Native American | 2 (0.2) | 1 (0.1) | 3 (0.1) | 0 (0.0) |
| Other | 5 (0.4) | 5 (0.3) | 9 (0.4) | 0. (0.0) |
| Pacific Islander | 0 (0.0) | 0 (0.0) | 1 (<0.01) | 0 (0.0) |
| White | 694 (57.8) | 1061 (57.5) | 1292 (52.8) | 0 (0.0) |

\*\*453 (96.2%) participants reported their ethnicity as Han Chinese, 18 (3.8%) participants reported their ethnicity as “other”

In the cohort serving through the pandemic onset (2019-2020 U.S. interns), there was a significant decrease in PHQ-9 depressive symptom scores from the pre-pandemic (September, December) to the pandemic period (April, May, June) (5.5 [3.9] vs. 4.9 [4.3]; *p*<0.001) period (total effect=-0.44, 95% CI [-0.61, -0.28]). Work hours also decreased significantly from pre-pandemic to the pandemic period (mean = 61.7 [standard deviation =14.4] hours/week pre-pandemic vs. 49.7 [20.7] hours/week during the pandemic, *p*<0.001).

Causal mediation analysis identified an average causally mediated effect (ACME) of -0.28 (95% CI [-0.34, -0.22]) for work hours, corresponding to 62% of the total effect on depressive symptoms being mediated through work hours (0.62, 95% CI [0.44-1.00]). After accounting for work hours, the difference in depressive symptom score between the pre-COVID-19 and COVID-19 assessments was attenuated (−0.17, 95% CI [-0.33, 0.00], Figure 1).

**Figure 1.**
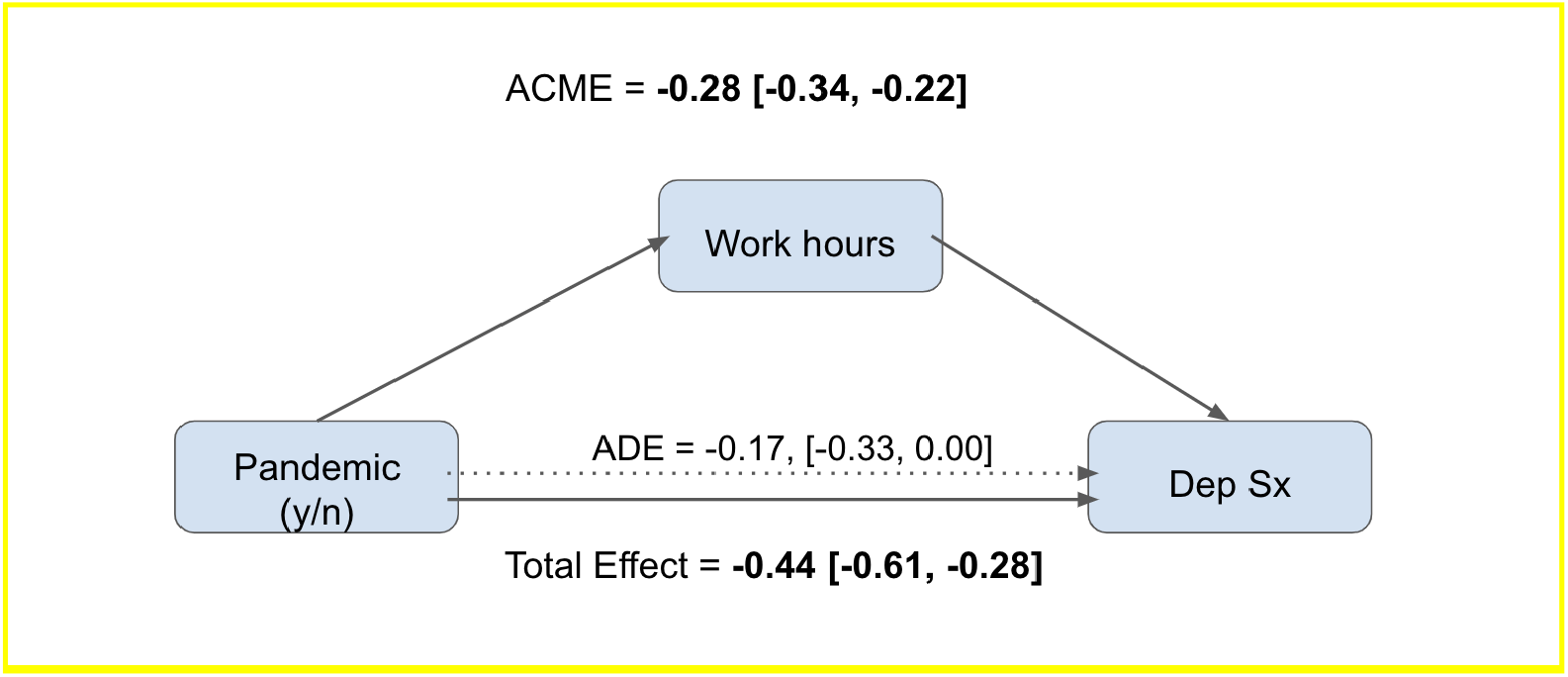
Causal mediation diagram showing the total effect, average direct effect (ADE), and average causally mediated effect of work hours on depressive symptoms within the 2019-20 cohort.

To further contextualize the changes in work hours and depressive symptoms observed in the 2019-2020 U.S. cohort, we descriptively compared this cohort’s average work hours and PHQ-9 scores to the parallel assessments from the 2018-2019 U.S. and 2020-2021 U.S. cohorts (Figure 2). Both work hours and depressive symptoms were substantially lower in spring of 2020 than in pre-pandemic cohorts. Further, both work hours and depressive symptoms nearly returned to pre-pandemic levels by fall of 2020.

**Figure 2.**
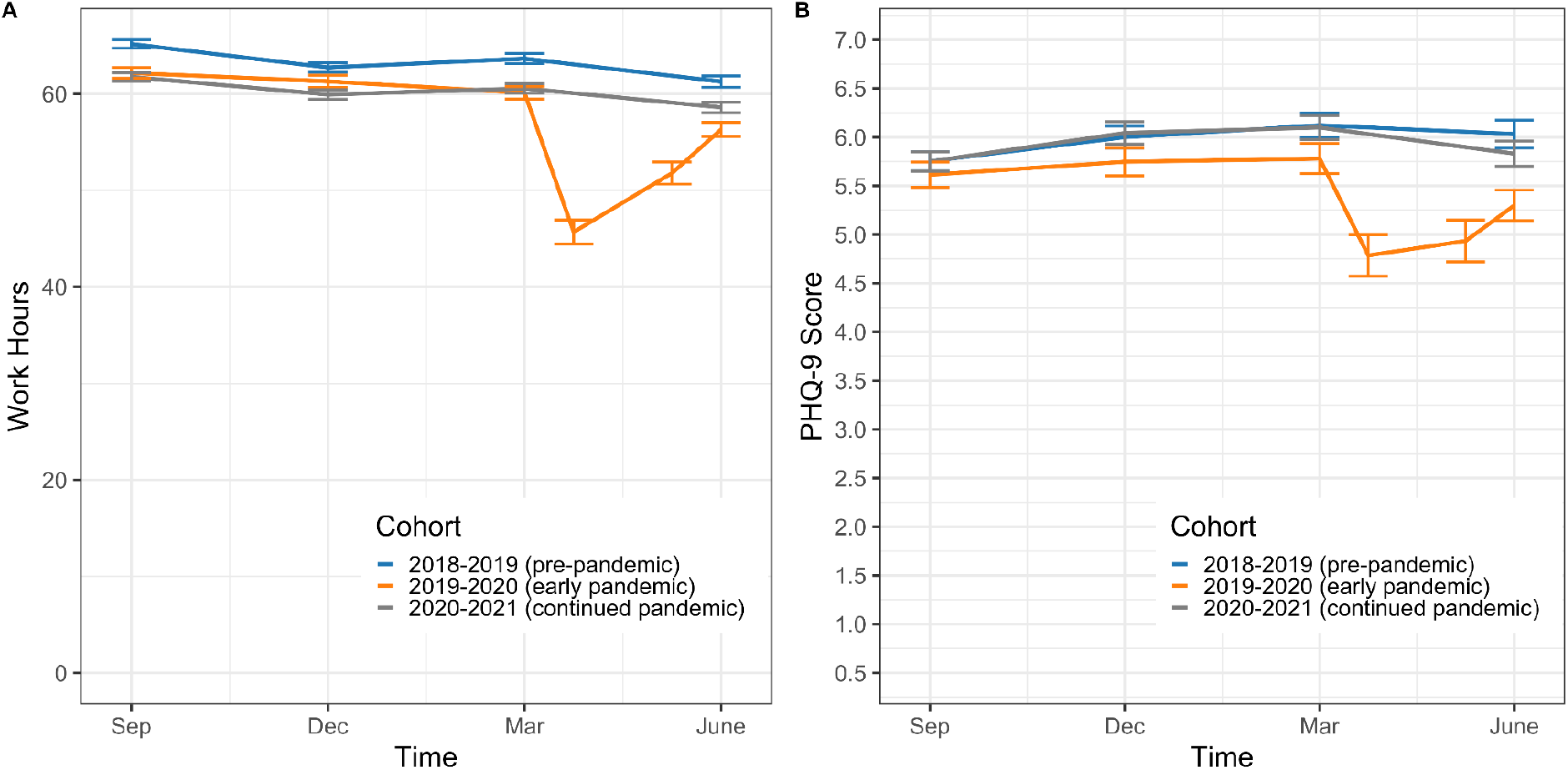
Work hours (A) and depressive symptoms (B) in the 2018-2019, 2019-2020, and 2020-2021 U.S. cohorts of the Intern Health Study. The 2019-2020 U.S. cohort demonstrated a drop in both work hours and depressive symptoms with the onset of the COVID-19 pandemic in Spring 2020.

**Figure 3.**
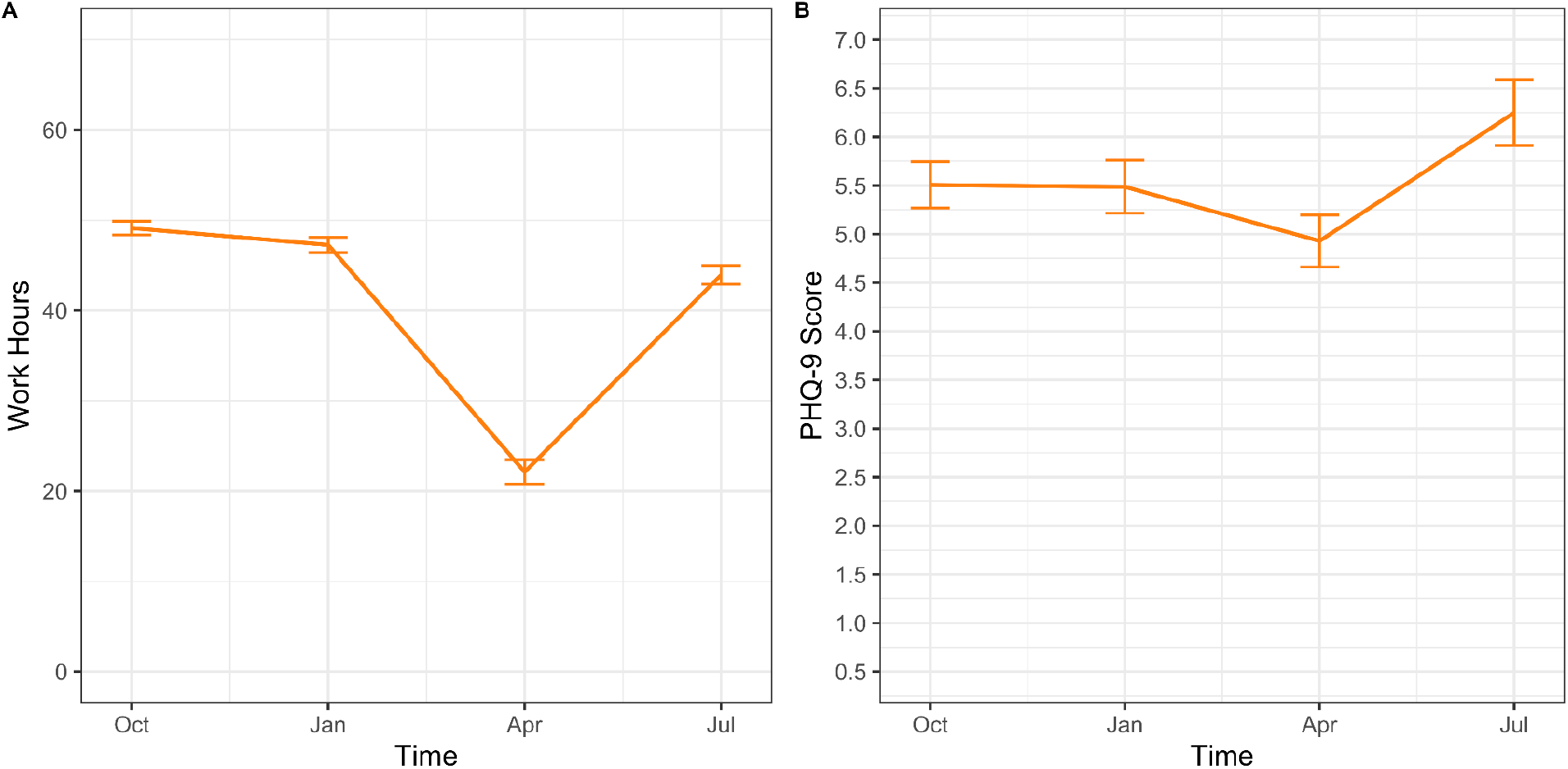
Work hours (A) and depressive symptoms (B) in the 2021-2022 Shanghai, China cohort of the Intern Health Study. This cohort demonstrated a drop in both work hours and depressive symptoms during the April 2022 Shanghai lockdown.

In the 2021-22 Shanghai cohort, depression scores decreased significantly between the pre-lockdown (October 2021, January 2022) and lockdown periods (April 2022) (5.56 [3.35] vs. 4.93 [4.8], p=0.005). After lockdown ended (July 2022), depression scores increased from the lockdown period (depression; 6.25 [5.4] vs. 4.93 [4.80], p<0.001) and were comparable to pre-lockdown levels (p’s> .01). Causal mediation analysis found an ACME of -0.40 [-0.63, -0.17] for work hours, indicating that 64.5% (0.64, 95% CI [0.24, 1.84]) of the total effect of lockdown on depressive symptoms was mediated through work hours. After accounting for work hours, the difference in depressive symptom scores between lockdown and non-lockdown assessments was attenuated (−0.22 [-0.67, 0.24]).

## DISCUSSION

Utilizing longitudinal cohorts of interns across the U.S. and Shanghai, China, we report the first systematic assessment of the trajectory of depressive symptoms in training physicians working before, during, and after the onset of the COVID-19 pandemic. Notably, U.S. interns experienced a 10.9% decrease in depressive symptoms during the early pandemic period, reporting the lowest PHQ-9 depressive symptom scores in the 16-year history of the Intern Health Study^18^. Similarly, interns training in Shanghai experienced an 11.3% decrease in depressive symptoms during the lockdown period. While moderate in magnitude, these decreases in depressive symptoms among interns are particularly noteworthy in the context of increasing depressive symptom burden in the general population during the COVID-19 pandemic^19,20^.

Our findings indicate that reduced workload is a potential explanation for this unexpected finding, as the majority of the decrease in depressive symptoms among residents in both the U.S. and Shanghai cohorts was mediated by a decrease in work hours in our study. U.S. residents reported working an average of 49.7 hours per week during the early pandemic period, corresponding to an average decrease of 12 hours from pre-pandemic levels. Similarly, Shanghai residents experienced a significant reduction in work hours during the 2022 lockdown, which was the primary driver of the 11.3% decrease in depressive symptoms. Prior studies suggest that work hours are a strong driver of depressive symptoms in resident physicians^15^. Our results add to this literature by demonstrating that duty hours mediated 62% of the decrease in depressive symptoms observed among U.S. interns during the pandemic onset and 64.5% of the total effect in Shanghai during the 2022 lockdown. However, in our analyses, we found that work hours nearly returned to pre-pandemic levels after the initial pandemic months, with a corresponding reversion of depressive symptoms to pre-pandemic levels. Similarly, results from the replication analysis in the Shanghai cohort demonstrated a reversion of both depressive symptoms and work hours to pre-lockdown levels in the months following the 2022 COVID-19 lockdown. These findings provide further evidence supporting the close link between work hours and depressive symptoms in training physicians.

This study has limitations. First, our sample consisted of intern physicians. Although the participating interns represent diverse demographic characteristics, experiences, and training environments, these findings may not generalize to other groups of physicians. Second, this study focused exclusively on depressive symptoms. It is possible that other mental health symptoms, such as anxiety symptoms, followed different trajectories through the pandemic. Finally, although residency programs, institutions and regulatory agencies implemented reforms specifically to reduce administrative and regulatory workload, we were unable to determine which specific activities contributed to the reduction in work hours during the early pandemic period.

With the threat of COVID-19 to their own health and the health of their patients, colleagues, and loved ones, the lives of training physicians were altered in unprecedented ways with the pandemic. In light of these changes, we identified a surprising decrease in depressive symptoms during the pandemic. This decrease, and the degree to which reduced work hours may be a driver of improved mental health, can inform measures to protect physician mental health and improve training physician well-being over the long-term.

## Data Availability

Data underlying the present analysis are available upon request to the authors, and data from the Intern Health Study are available from ICPSR.

## Disclaimer

The opinions, results, and conclusions reported in this article are those of the authors and are independent from the funding sources.

